# Who’s Left Behind? Exploring the Digital Divide in Health App Use Among Adults with Hypertension and Diabetes

**DOI:** 10.1101/2025.11.24.25340934

**Authors:** Nana Ofori Adomako, Kezia Owusu Nti, Joyline Chepkorir, Kwabena Asabere Asante, Ibrahim Dehshini Ali, Nicholas Ofori, Ethel Emefa Agordekpe, Yuling Chen, Hailey N Miller, Cheryl R Himmelfarb

## Abstract

**Background:** Mobile health (mHealth) applications are promising tools for managing chronic conditions such as hypertension and diabetes. However, disparities in access and use, often described as the digital divide, may limit their benefits for vulnerable populations

**Objective:** To assess the prevalence of mHealth app use among U.S. adults with hypertension or diabetes and examine sociodemographic, technological, and health-related factors associated with digital engagement.

**Methods:** Data were drawn from the 2024 Health Information National Trends Survey (HINTS 7). Respondents with self-reported hypertension or diabetes (n=3,172) were included. The outcome was self-reported mHealth app use. Independent variables included sociodemographic characteristics, technological access and health-related factors. Survey-weighted log-binomial regression estimated adjusted prevalence ratios (RRs) and 95% confidence intervals (CIs).

**Results:** Significant disparities in mHealth app use were observed. Adults aged 65–74 (RR=0.70, 95% CI: 0.56–0.88) and 75+ years (RR=0.50, 95% CI: 0.35–0.71) were less likely to use apps than younger adults. Males (RR=0.85, 95% CI: 0.74–0.97) reported lower use than females.

Higher income (≥$100,000: RR=1.39, 95% CI: 1.06–1.81) and education (college+: RR=1.42, 95% CI: 1.17–1.73) were associated with greater use. Smartphone non-ownership (RR=0.42, 95% CI: 0.24–0.75), never using the internet (RR=0.07, 95% CI: 0.01–0.46), lower digital confidence (RR=0.74, 95% CI: 0.56–0.97), and high frustration with technology (RR=0.61, 95% CI: 0.43–0.87) were strong barriers. Health-related factors were not significantly associated.

**Conclusion:** Persistent disparities in mHealth use among adults with hypertension and diabetes underscore the need to address digital access and literacy to promote equitable engagement with digital health tools.

## Introduction

Hypertension and diabetes are among the most prevalent chronic conditions globally, contributing substantially to morbidity, mortality, and healthcare costs [1]. In the United States, an estimated 129 million adults live with at least one major chronic disease, with diabetes alone affecting over 38 million adults as of 2025 [2]. These conditions disproportionately impact older adults, racially and ethnically minoritized populations, and individuals with lower socioeconomic status that also face systemic barriers to care such as limited insurance coverage, geographic isolation, and reduced access to culturally responsive services [3]. Effective management hinges on consistent medication adherence, lifestyle modifications, and timely monitoring, yet many patients struggle with fragmented care, low health literacy, and high treatment burden [4].

Despite growing interest in digital health solutions, few studies have examined mHealth app use specifically among adults with hypertension or diabetes using nationally representative data.

Much of the existing literature focuses on general populations or restricted clinical samples, limiting the generalizability of findings [5–7].

Mobile health (mHealth) applications (apps) have become increasingly important in chronic disease management, providing functionalities such as medication tracking, symptom monitoring, and personalized health education [8]. For individuals living with hypertension and diabetes, conditions that require ongoing self-management and self-monitoring, these tools can strengthen patient engagement, improve adherence, and support lifestyle modifications [9]. The widespread adoption of smartphones and digital platforms has created new opportunities for scalable and cost-effective interventions that extend beyond and complement traditional clinical settings [10]

Despite these opportunities, the benefits of mHealth are not equitably distributed. The digital divide, encompassing disparities in access to technology, internet connectivity, and digital literacy, remains a significant barrier to health equity [11,12]. Older adults, individuals with lower income or educational attainment, and racially and ethnically minoritized populations are disproportionately affected by limited digital access and confidence in using technology [13,14]. These disparities are particularly concerning because these groups also experience higher burdens of chronic disease and persistent barriers to care [15].

Digital literacy, defined as the ability to effectively navigate, evaluate, and use digital tools, is increasingly recognized as a social determinant of health [16]. Limited digital literacy can lead to frustration, mistrust, and disengagement from digital health platforms, undermining their intended benefits [17]. Moreover, the intersection of technological barriers with broader structural inequities, including language barriers, geographic isolation, and historical mistrust, can further compound disparities in mHealth engagement [13,14].

While digital health equity continues to gain momentum, there remains a notable gap in research focused on mHealth app use among individuals with hypertension and diabetes, particularly studies grounded in nationally representative data. Existing evidence often centers on general or narrowly defined clinical populations, which constrains our understanding of how these tools are adopted and experienced across diverse, real-world settings. There is also a need to disentangle the relative contributions of sociodemographic, technological, and psychosocial factors to digital engagement in chronic disease management.

Understanding how sociodemographic, technological, and psychosocial factors jointly influence mHealth engagement is critical to designing inclusive digital health interventions. To address these gaps, this study analyses data from the 2024 Health Information National Trends Survey (HINTS 7), focusing on U.S. adults with self-reported hypertension or diabetes [20]. Our objectives are to assess the prevalence of mHealth app use and to examine factors associated with digital engagement, with particular attention to disparities in access, literacy, and confidence.

## Methods

### Study Design and Data Source

This cross-sectional study utilized data from the 2024 Health Information National Trends Survey (HINTS 7), a nationally representative survey administered by the National Cancer Institute [20]. HINTS collects data on health-related information-seeking behaviors, digital health engagement, and chronic disease management among U.S. adults. The survey employs a stratified, two-stage sampling design to ensure representation across sociodemographic groups, including oversampling of Hispanic and non-Hispanic Black populations.

### Study Population

The analytic sample included adults aged 18 years and older who self-reported a physician diagnosis of hypertension and/or diabetes. Participants were eligible if they responded to questions regarding mobile health application use. Individuals with missing data on chronic disease (diabetes and hypertension) status or mobile health application use were excluded using listwise deletion. The final analytic sample comprised 3,172 adults with diabetes or hypertension. All analyses incorporated HINTS 7 sampling weights and design variables to produce nationally representative estimates. The sample selection process is detailed in the figure below (Figure 1).

**Figure 1:**
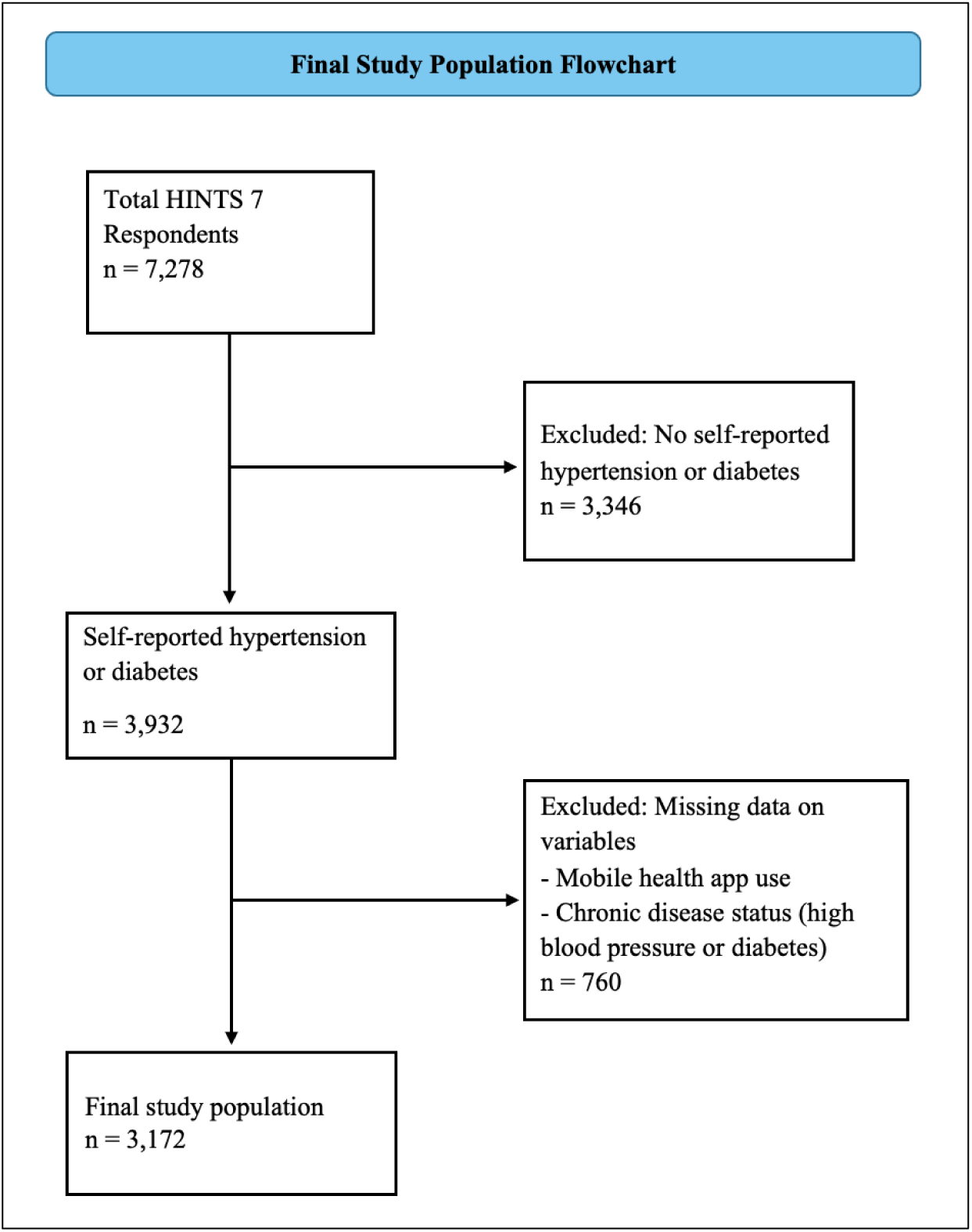
Flow Diagram of Sample Selection from HINTS 7 (2024)

### Outcome Variable

The primary outcome was self-reported use of mobile health applications for health-related purposes [21]. Participants were asked “In the past 12 months, have you used a health or wellness app on your tablet or smartphone?” Responses were dichotomized (Yes/No), consistent with prior analysis using the HINTS dataset [21].

### Independent Variables

Independent variables were grouped into three domains:

- Sociodemographic characteristics included age (categorized as 18–34, 35–49, 50–64, 65–74, 75+), sex, race/ethnicity (non-Hispanic White, non-Hispanic Black, Hispanic, non-Hispanic Asian, American Indian or Alaska Native, Native Hawaiian or Other Pacific Islander, some other race, and multiracial), education level (high school to postgraduate), annual household income ($0–$200k+), and rural vs. urban residence.
- Technological access and use: smartphone ownership (yes/no), frequency of internet use (daily, weekly, rarely, never).

Digital literacy and confidence: Digital literacy and confidence were assessed using self-reported Likert-scale items from the HINTS 7 questionnaire. Participants rated their agreement (Strongly agree to Strongly disagree) with the following statements:

- *“I have the skills to find the health information I need on the Internet.”*
- *“I can use applications/programs (like Zoom) on my cell phone or computer without asking someone for help.”*
- *“I find learning how to use new technology frustrating.”*

Responses were categorized to reflect levels of digital self-efficacy, independence, and frustration. These items were used to construct composite indicators of digital literacy and confidence, consistent with prior HINTS-based studies [22].

### Health-Related Covariates

Health-related factors were derived from self-reported items in the HINTS 7 questionnaire and included:

- Number of chronic conditions: Participants indicated whether they had ever been diagnosed with specific conditions (e.g., diabetes, hypertension, heart disease). Responses were summed to create a count variable.
- Smoking status: Assessed via items on current and past tobacco use (e.g., “Do you now smoke cigarettes every day, some days, or not at all?”).
- Alcohol use: Measured by frequency of alcohol consumption in the past 30 days.
- Physical activity: Based on self-reported frequency of moderate or vigorous exercise per week.
- Sleep quality: Evaluated using the item “In general, how would you rate your sleep quality?” with responses ranging from “Very good” to “Very bad.”

Detailed question wording, response options, and coding procedures are provided in the Supplemental Materials.

## Statistical Analysis

Survey weights provided by HINTS were applied to account for the complex sampling design and ensure national representativeness. Descriptive statistics were used to characterize the sample. Bivariate analyses were conducted using chi-square tests and t-tests, as appropriate.

Multivariable log-binomial regression models were used to estimate adjusted prevalence ratios (aPRs) and 95% confidence intervals (CIs) for mobile health app use. Covariates included:

- Sociodemographic characteristics: age, sex, race/ethnicity, education level, household income, and rural vs. urban residence
- Technological access and use: smartphone ownership and frequency of internet use
- Digital literacy and confidence: self-reported confidence using digital devices, frustration with technology, and perceived internet search skills
- Health-related factors: number of chronic conditions, smoking status, alcohol use, physical activity, and sleep quality

Covariates were selected based on theoretical relevance, prior literature on digital health engagement, and availability within the HINTS 7 dataset. All analyses were conducted using R version 4.3.1, and statistical significance was defined as a two-sided p-value < 0.05.[23].

## Results

### Sociodemographic Characteristics by mHealth Use

Table 1 presents the baseline characteristics of users and non-users of mobile health apps with hypertension or diabetes. Compared with non-users, users were more likely to be younger (ages 18–49), have higher educational attainment, and report higher household income (all p < 0.001). Users reported higher levels of internet use, greater confidence in search skills, and increased ownership of digital devices including smartphones, tablets, and smartwatches (all p < 0.001).

**Table 1:**
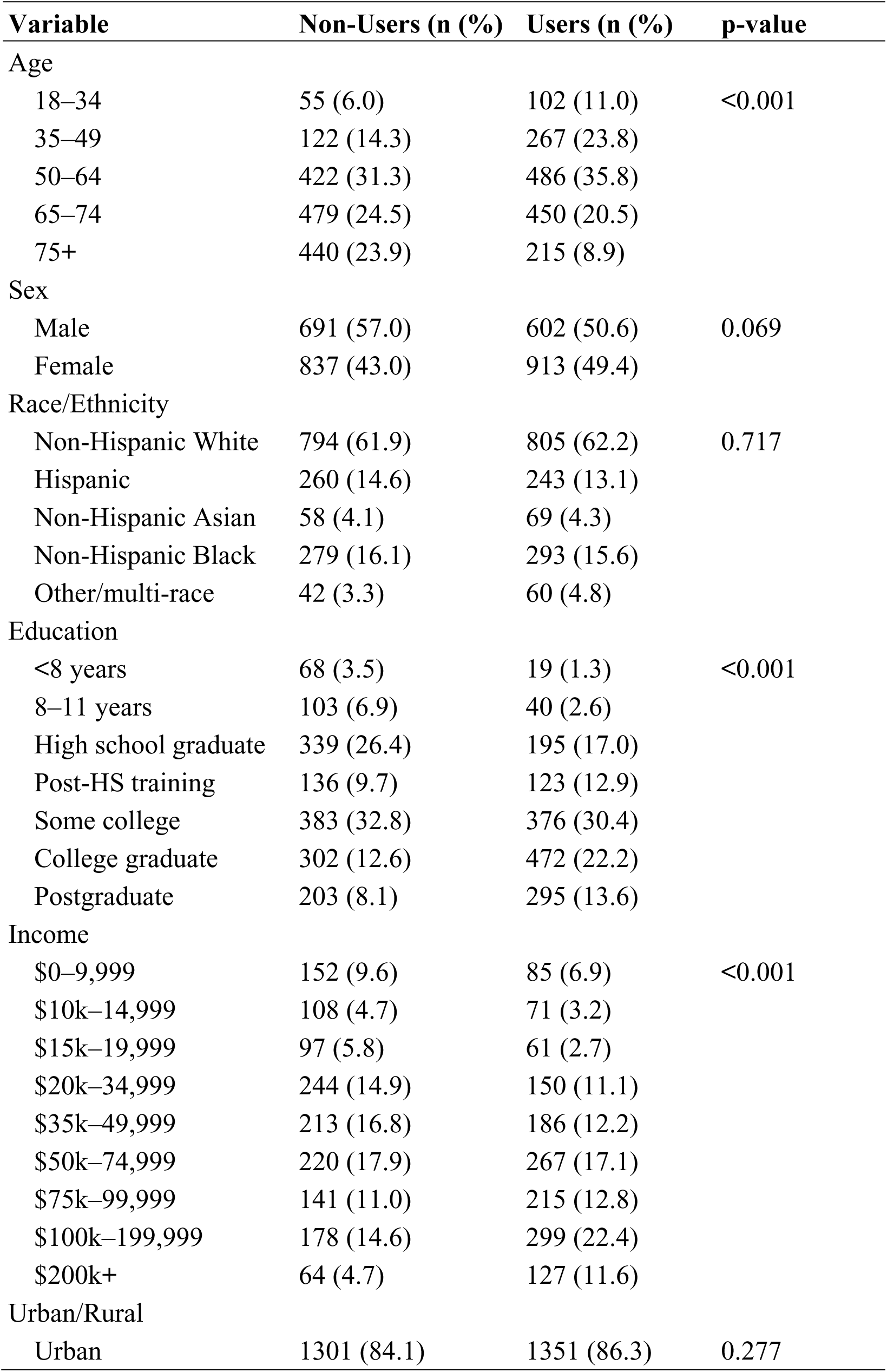
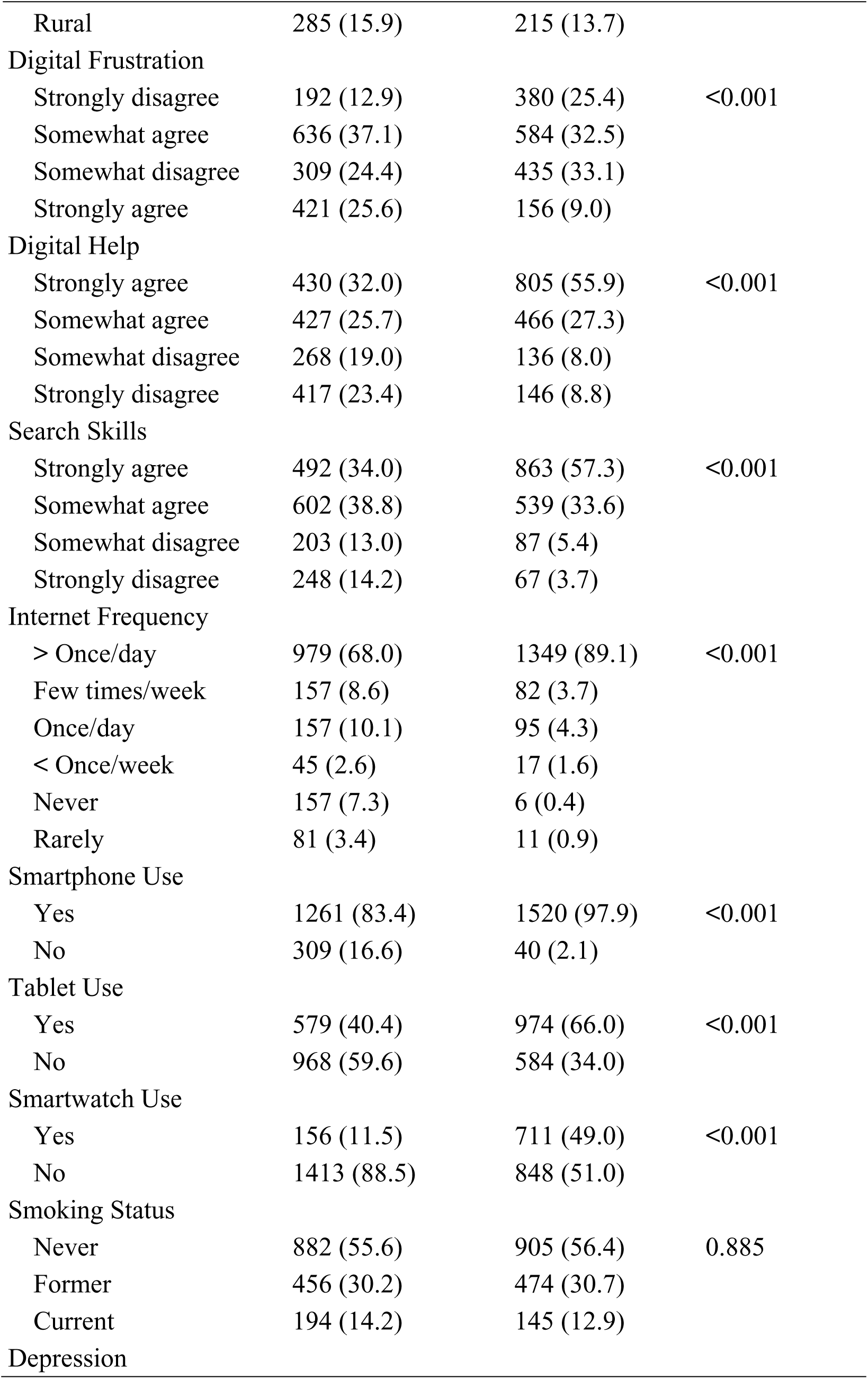
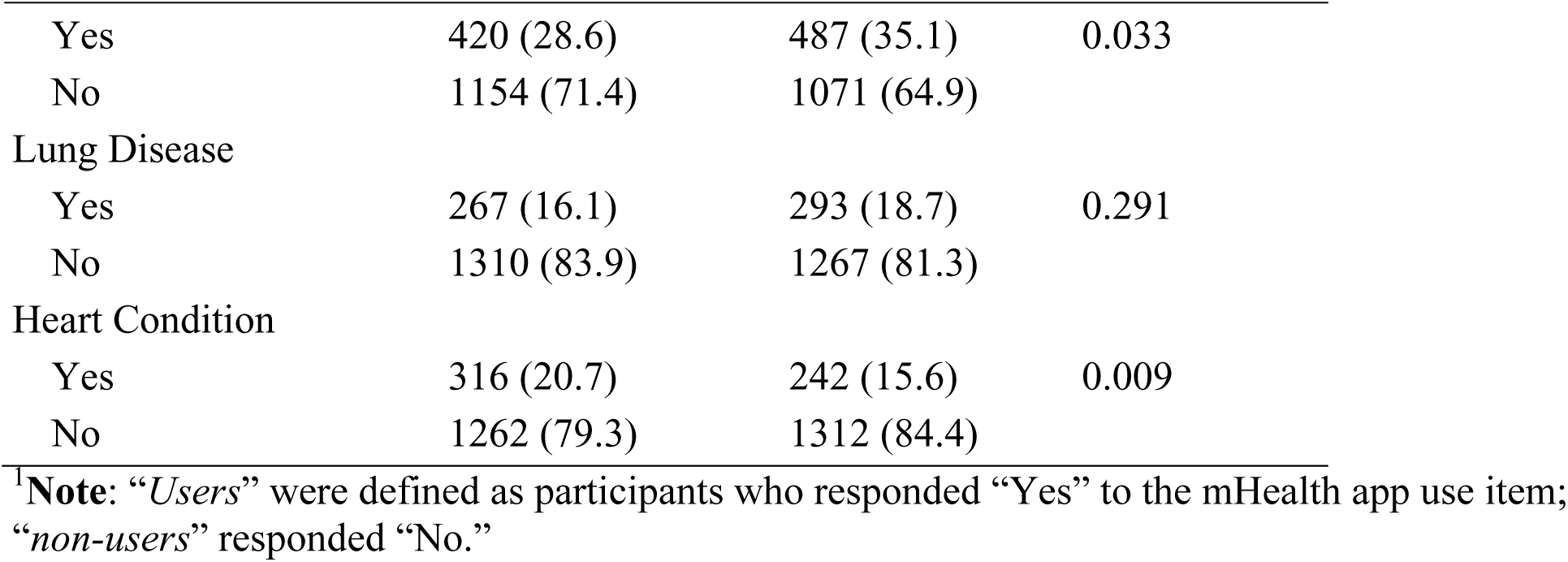
Socio-demographic characteristics of U.S. adults with hypertension or diabetes, stratified by mHealth app use (N=3,172)

| Variable | Non-Users (n (%)) | Users (n (%)) | p-value |
| --- | --- | --- | --- |
| Age |  |  |  |
| 18–34 | 55 (6.0) | 102 (11.0) | <0.001 |
| 35–49 | 122 (14.3) | 267 (23.8) |  |
| 50–64 | 422 (31.3) | 486 (35.8) |  |
| 65–74 | 479 (24.5) | 450 (20.5) |  |
| 75+ | 440 (23.9) | 215 (8.9) |  |
| Sex |  |  |  |
| Male | 691 (57.0) | 602 (50.6) | 0.069 |
| Female | 837 (43.0) | 913 (49.4) |  |
| Race/Ethnicity |  |  |  |
| Non-Hispanic White | 794 (61.9) | 805 (62.2) | 0.717 |
| Hispanic | 260 (14.6) | 243 (13.1) |  |
| Non-Hispanic Asian | 58 (4.1) | 69 (4.3) |  |
| Non-Hispanic Black | 279 (16.1) | 293 (15.6) |  |
| Other/multi-race | 42 (3.3) | 60 (4.8) |  |
| Education |  |  |  |
| <8 years | 68 (3.5) | 19 (1.3) | <0.001 |
| 8–11 years | 103 (6.9) | 40 (2.6) |  |
| High school graduate | 339 (26.4) | 195 (17.0) |  |
| Post-HS training | 136 (9.7) | 123 (12.9) |  |
| Some college | 383 (32.8) | 376 (30.4) |  |
| College graduate | 302 (12.6) | 472 (22.2) |  |
| Postgraduate | 203 (8.1) | 295 (13.6) |  |
| Income |  |  |  |
| \$0–9,999 | 152 (9.6) | 85 (6.9) | <0.001 |
| \$10k–14,999 | 108 (4.7) | 71 (3.2) | |
| \$15k–19,999 | 97 (5.8) | 61 (2.7) | |
| \$20k–34,999 | 244 (14.9) | 150 (11.1) | |
| \$35k–49,999 | 213 (16.8) | 186 (12.2) | |
| \$50k–74,999 | 220 (17.9) | 267 (17.1) | |
| \$75k–99,999 | 141 (11.0) | 215 (12.8) | |
| \$100k–199,999 | 178 (14.6) | 299 (22.4) | |
| \$200k+ | 64 (4.7) | 127 (11.6) | |
| Urban/Rural |  |  |  |
| Urban | 1301 (84.1) | 1351 (86.3) | 0.277 |

| **Variable** | **Non-Users (n (%))** | **Users (n (%))** | **p-value** |
| --- | --- | --- | --- |
| Rural | 285 (15.9) | 215 (13.7) |  |
| Digital Frustration |  |  |  |
| Strongly disagree | 192 (12.9) | 380 (25.4) | <0.001 |
| Somewhat agree | 636 (37.1) | 584 (32.5) |  |
| Somewhat disagree | 309 (24.4) | 435 (33.1) |  |
| Strongly agree | 421 (25.6) | 156 (9.0) |  |
| Digital Help |  |  |  |
| Strongly agree | 430 (32.0) | 805 (55.9) | <0.001 |
| Somewhat agree | 427 (25.7) | 466 (27.3) |  |
| Somewhat disagree | 268 (19.0) | 136 (8.0) |  |
| Strongly disagree | 417 (23.4) | 146 (8.8) |  |
| Search Skills |  |  |  |
| Strongly agree | 492 (34.0) | 863 (57.3) | <0.001 |
| Somewhat agree | 602 (38.8) | 539 (33.6) |  |
| Somewhat disagree | 203 (13.0) | 87 (5.4) |  |
| Strongly disagree | 248 (14.2) | 67 (3.7) |  |
| Internet Frequency |  |  |  |
| > Once/day | 979 (68.0) | 1349 (89.1) | <0.001 |
| Few times/week | 157 (8.6) | 82 (3.7) |  |
| Once/day | 157 (10.1) | 95 (4.3) |  |
| < Once/week | 45 (2.6) | 17 (1.6) |  |
| Never | 157 (7.3) | 6 (0.4) |  |
| Rarely | 81 (3.4) | 11 (0.9) |  |
| Smartphone Use |  |  |  |
| Yes | 1261 (83.4) | 1520 (97.9) | <0.001 |
| No | 309 (16.6) | 40 (2.1) |  |
| Tablet Use |  |  |  |
| Yes | 579 (40.4) | 974 (66.0) | <0.001 |
| No | 968 (59.6) | 584 (34.0) |  |
| Smartwatch Use |  |  |  |
| Yes | 156 (11.5) | 711 (49.0) | <0.001 |
| No | 1413 (88.5) | 848 (51.0) |  |
| Smoking Status |  |  |  |
| Never | 882 (55.6) | 905 (56.4) | 0.885 |
| Former | 456 (30.2) | 474 (30.7) |  |
| Current | 194 (14.2) | 145 (12.9) |  |
| Depression |  |  |  |
| Yes | 420 (28.6) | 487 (35.1) | 0.033 |
| No | 1154 (71.4) | 1071 (64.9) |  |
| Lung Disease |  |  |  |
| Yes | 267 (16.1) | 293 (18.7) | 0.291 |
| No | 1310 (83.9) | 1267 (81.3) |  |
| Heart Condition |  |  |  |
| Yes | 316 (20.7) | 242 (15.6) | 0.009 |
| No | 1262 (79.3) | 1312 (84.4) |  |

<sup>†</sup>**Note:** “Users” were defined as participants who responded “Yes” to the mHealth app use item; “non-users” responded “No.”

Psychosocial and clinical differences were also observed. Users reported a higher prevalence of depression (p = 0.033) and a lower prevalence of heart conditions (p = 0.009). No statistically significant differences were found by sex, race/ethnicity, smoking status, or lung disease (Table 1).

### Socio-demographic Differences in mHealth App use

In bivariate regression models, age was significantly associated with mHealth app use, with older adults demonstrating markedly lower engagement compared to younger counterparts.

Specifically, individuals aged 65–74 (RR = 0.70, 95% CI: 0.56–0.89, p = 0.004) and 75+ (RR = 0.50, 95% CI: 0.35–0.70, p < 0.001) had significantly reduced likelihoods of app use compared to those aged 18–34. Racial and ethnic differences were not statistically significant, although Hispanic (RR = 0.91, 95% CI: 0.75–1.11, p = 0.350) and Non-Hispanic Asian adults (RR = 0.84, 95% CI: 0.63–1.12, p = 0.226) exhibited slightly lower usage rates. Socioeconomic factors showed strong positive associations: adults with annual incomes of $100,000–199,999 (RR = 1.19, 95% CI: 1.02–1.38, p = 0.027) and $200,000+ (RR = 1.24, 95% CI: 1.03–1.49, p = 0.027) were significantly more likely to use mHealth apps compared to those earning $50,000–74,999. Educational attainment was also positively associated with app engagement. Compared to high school graduates, individuals with vocational/technical training (RR = 1.46, 95% CI: 1.14–1.86, p = 0.004), college degrees (RR = 1.37, 95% CI: 1.10–1.71, p = 0.007), and postgraduate education (RR = 1.35, 95% CI: 1.04–1.75, p = 0.026) had significantly greater likelihoods of app use. No significant differences were observed between urban and rural residents (RR = 1.00, 95% CI: 0.84–1.18, p = 0.998). (Table 2, Figure 2)

**Figure 2:**
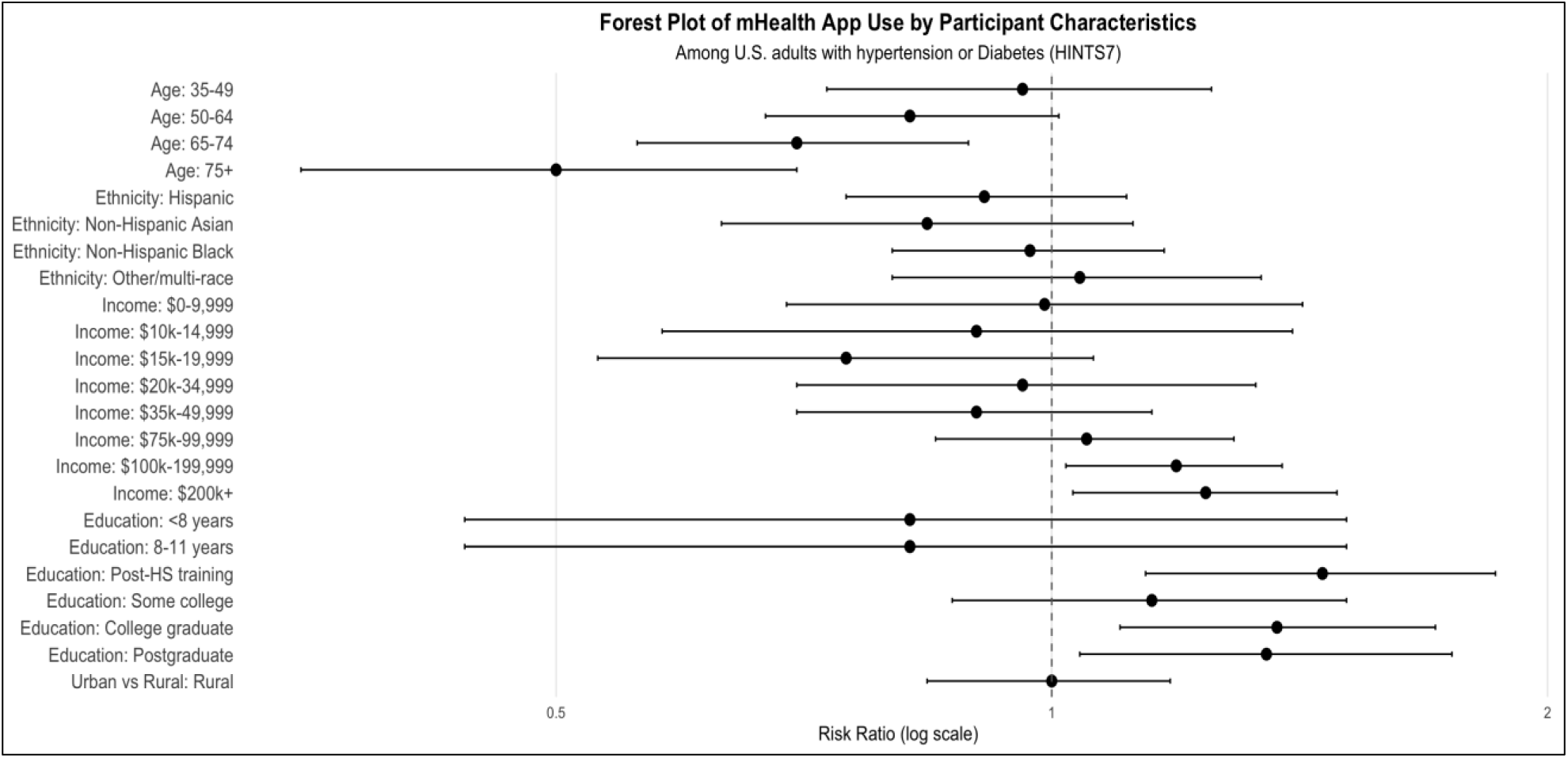
Forest plot of adjusted prevalence ratios (aPRs) for mobile health app use among U.S. adults with hypertension or diabetes, stratified by sociodemographic characteristics. Circles represent point estimates of aPRs, horizontal lines indicate 95% confidence intervals, and the vertical dashed line denotes the null value (RR=1). Reference categories include age 18–34 years, female sex, high school education, annual household income $50,000–74,999, and urban residence.

**Table 2:**
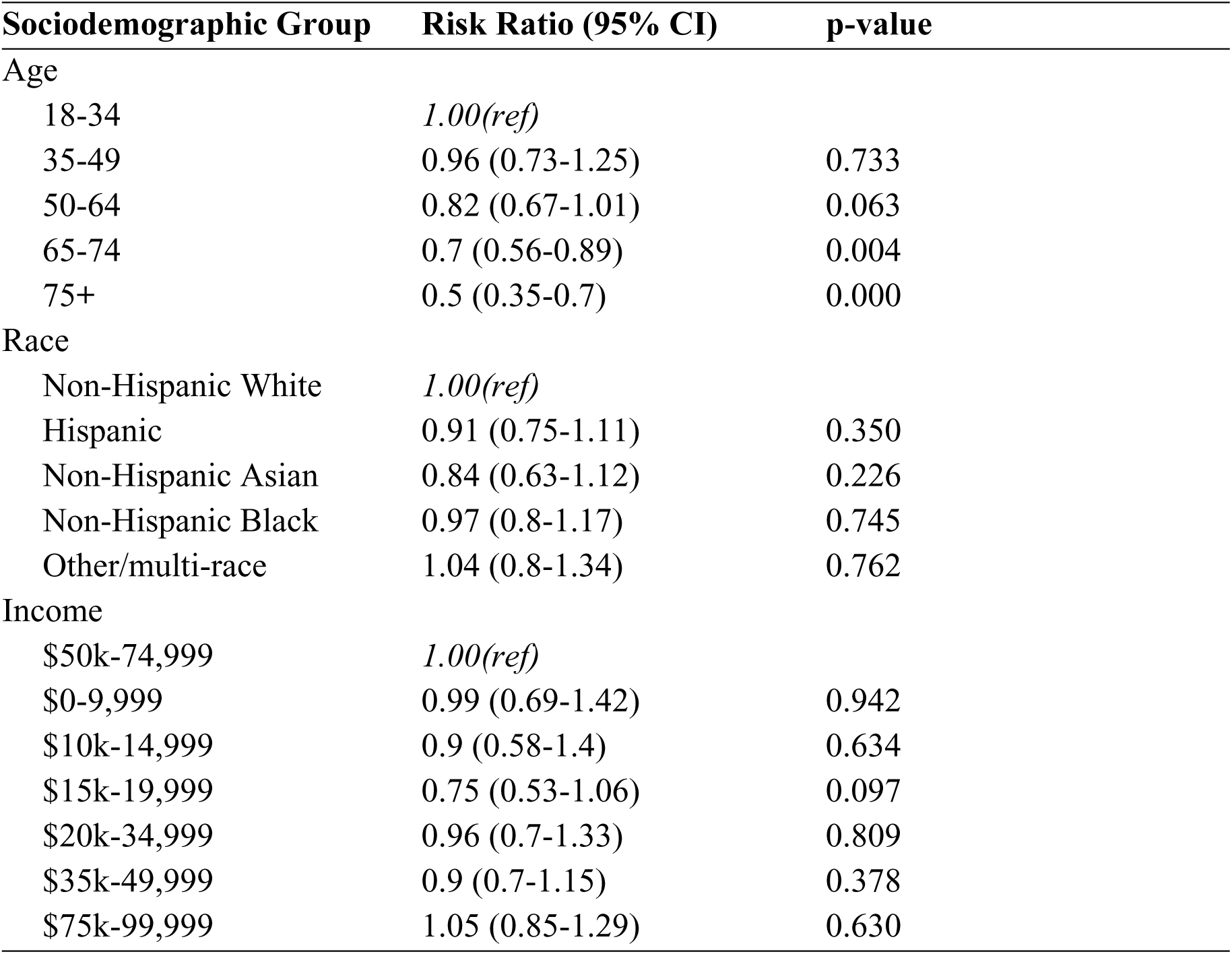
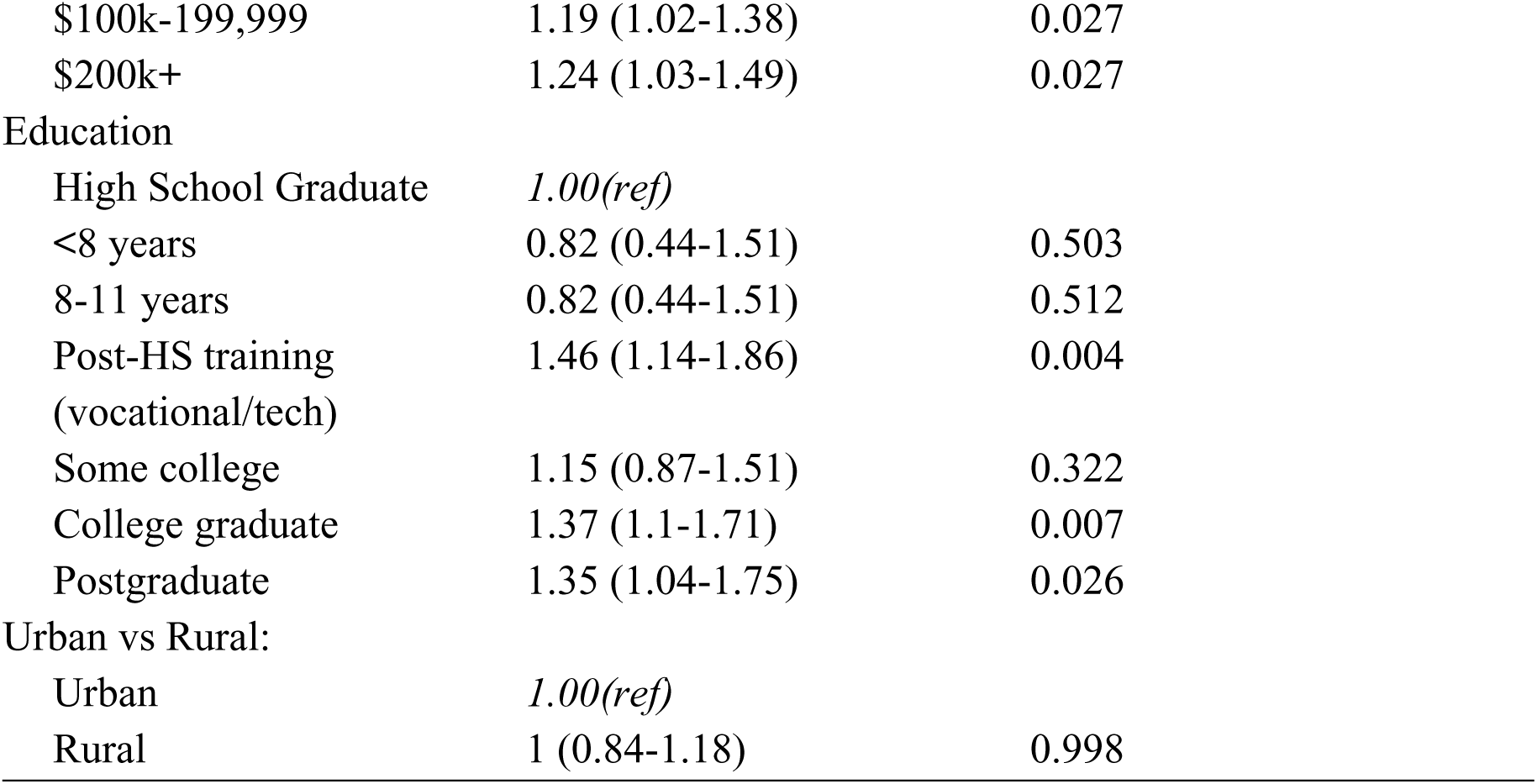
Bivariate Associations Between Participant Characteristics and mHealth App Use (HINTS 7)

| Sociodemographic Group | Risk Ratio (95% CI) | p-value |
| --- | --- | --- |
| Age |  |  |
| 18-34 | *1.00(ref)* |  |
| 35-49 | 0.96 (0.73-1.25) | 0.733 |
| 50-64 | 0.82 (0.67-1.01) | 0.063 |
| 65-74 | 0.7 (0.56-0.89) | 0.004 |
| 75+ | 0.5 (0.35-0.7) | 0.000 |
| Race |  |  |
| Non-Hispanic White | *1.00(ref)* |  |
| Hispanic | 0.91 (0.75-1.11) | 0.350 |
| Non-Hispanic Asian | 0.84 (0.63-1.12) | 0.226 |
| Non-Hispanic Black | 0.97 (0.8-1.17) | 0.745 |
| Other/multi-race | 1.04 (0.8-1.34) | 0.762 |
| Income |  |  |
| \$50k-74,999 | *1.00(ref)* | |
| \$0-9,999 | 0.99 (0.69-1.42) | 0.942 |
| \$10k-14,999 | 0.9 (0.58-1.4) | 0.634 |
| \$15k-19,999 | 0.75 (0.53-1.06) | 0.097 |
| \$20k-34,999 | 0.96 (0.7-1.33) | 0.809 |
| \$35k-49,999 | 0.9 (0.7-1.15) | 0.378 |
| \$75k-99,999 | 1.05 (0.85-1.29) | 0.630 |
| \$100k-199,999 | 1.19 (1.02-1.38) | 0.027 |
| \$200k+ | 1.24 (1.03-1.49) | 0.027 |
| Education |  |  |
| High School Graduate | *1.00(ref)* |  |
| <8 years | 0.82 (0.44-1.51) | 0.503 |
| 8-11 years | 0.82 (0.44-1.51) | 0.512 |
| Post-HS training<br>(vocational/tech) | 1.46 (1.14-1.86) | 0.004 |
| Some college | 1.15 (0.87-1.51) | 0.322 |
| College graduate | 1.37 (1.1-1.71) | 0.007 |
| Postgraduate | 1.35 (1.04-1.75) | 0.026 |
| Urban vs Rural: |  |  |
| Urban | *1.00(ref)* |  |
| Rural | 1 (0.84-1.18) | 0.998 |

### Technological Access and Confidence

Technological access and digital confidence were associated with mHealth app use among adults with hypertension or diabetes. Individuals without a smartphone were significantly less likely to engage with health apps (RR = 0.40, 95% CI: 0.23–0.69, p = 0.003), and those who never used the internet had dramatically reduced odds of use (RR = 0.06, 95% CI: 0.01–0.42, p = 0.008).

Beyond access, attitudes toward technology played a meaningful role: respondents who somewhat disagreed with the statement “I can use technology without help” were less likely to use apps (RR = 0.74, 95% CI: 0.56–0.97, p = 0.032), and those who strongly agreed with feeling frustrated by technology also showed reduced engagement (RR = 0.66, 95% CI: 0.49–0.90, p = 0.011). Although lower self-reported internet search skills were associated with decreased app use, these findings did not reach statistical significance. (Table 3, Figure 3).

**Figure 3:**
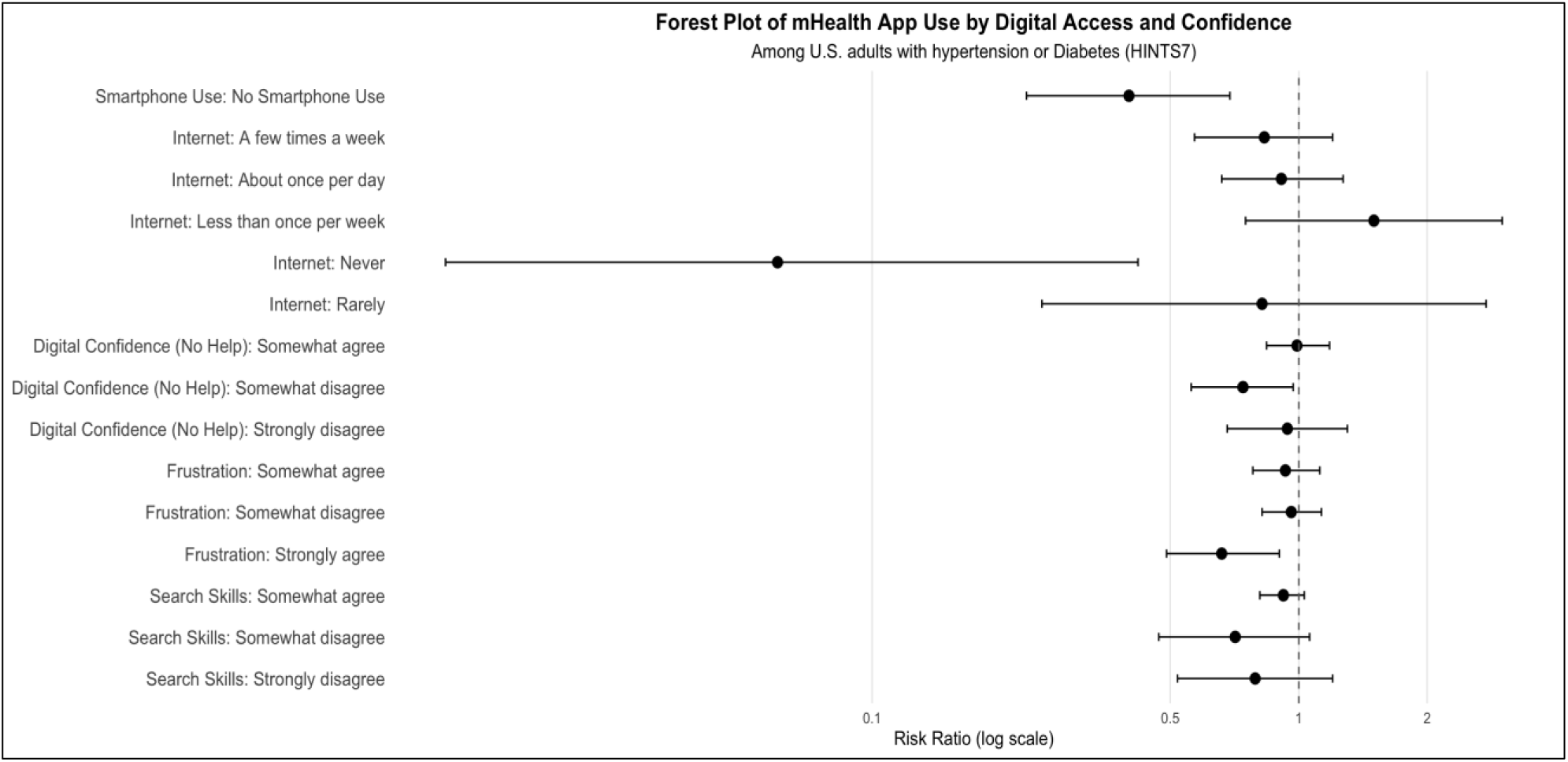
Forest plot of adjusted prevalence ratios (aPRs) for mobile health app use by technological access and digital confidence. Circles represent point estimates, horizontal lines indicate 95% confidence intervals, and the vertical dashed line denotes the null value (RR=1). Reference categories include smartphone ownership, daily internet use, high confidence in technology use, and low frustration with technology.

**Figure 4:**
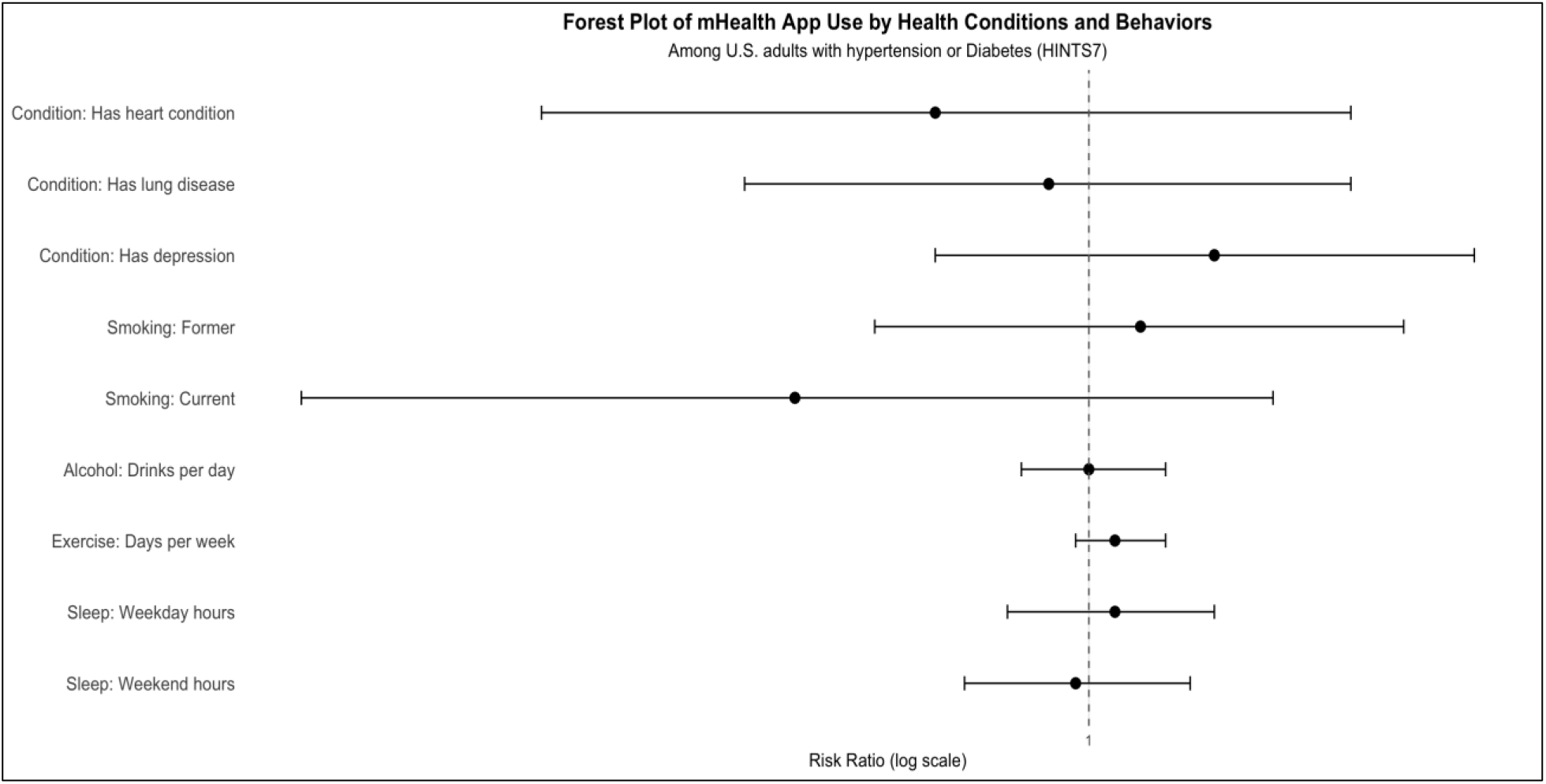
Forest plot of adjusted prevalence ratios (aPRs) for mobile health app use by health-related factors among U.S. adults with hypertension or diabetes. Circles represent point estimates, horizontal lines indicate 95% confidence intervals, and the vertical dashed line denotes the null value (RR=1). Reference categories include never smoker, absence of comorbid conditions, and average sleep quality. Continuous variables (e.g., number of chronic conditions, physical activity frequency) are modeled per unit increase.

**Table 3:**
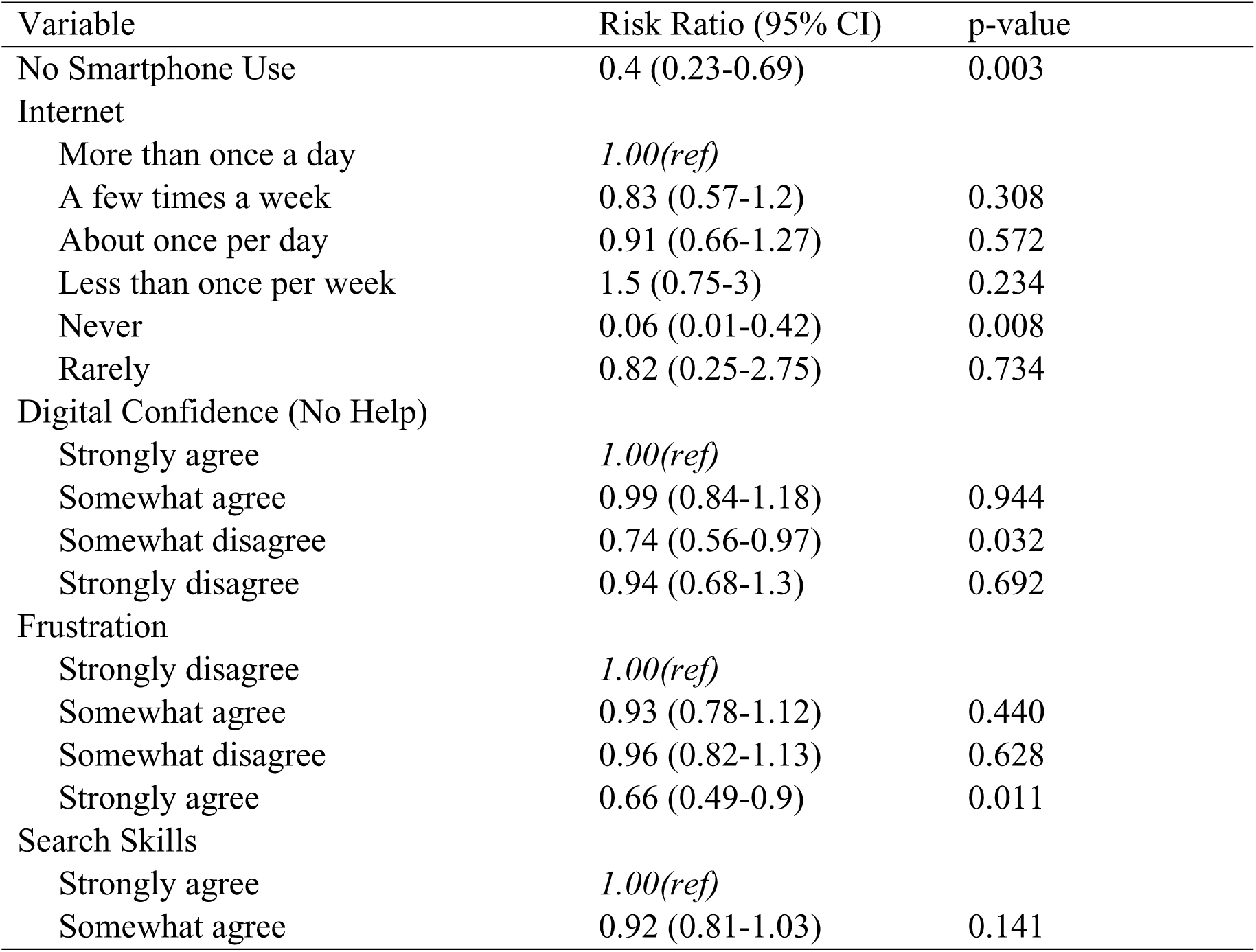
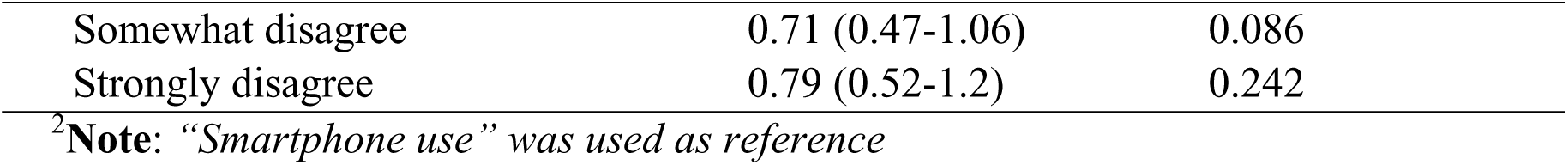
Adjusted risk ratios for health app use by technological factors among U.S. adults with hypertension or diabetes (HINTS 7)

| Variable | Risk Ratio (95% CI) | p-value |
| --- | --- | --- |
| No Smartphone Use | 0.4 (0.23-0.69) | 0.003 |
| Internet |  |  |
| More than once a day | *1.00(ref)* |  |
| A few times a week | 0.83 (0.57-1.2) | 0.308 |
| About once per day | 0.91 (0.66-1.27) | 0.572 |
| Less than once per week | 1.5 (0.75-3) | 0.234 |
| Never | 0.06 (0.01-0.42) | 0.008 |
| Rarely | 0.82 (0.25-2.75) | 0.734 |
| Digital Confidence (No Help) |  |  |
| Strongly agree | *1.00(ref)* |  |
| Somewhat agree | 0.99 (0.84-1.18) | 0.944 |
| Somewhat disagree | 0.74 (0.56-0.97) | 0.032 |
| Strongly disagree | 0.94 (0.68-1.3) | 0.692 |
| Frustration |  |  |
| Strongly disagree | *1.00(ref)* |  |
| Somewhat agree | 0.93 (0.78-1.12) | 0.440 |
| Somewhat disagree | 0.96 (0.82-1.13) | 0.628 |
| Strongly agree | 0.66 (0.49-0.9) | 0.011 |
| Search Skills |  |  |
| Strongly agree | *1.00(ref)* |  |
| Somewhat agree | 0.92 (0.81-1.03) | 0.141 |
| Somewhat disagree | 0.71 (0.47-1.06) | 0.086 |
| Strongly disagree | 0.79 (0.52-1.2) | 0.242 |

<sup>2</sup>**Note:** “*Smartphone use*” was used as reference

### Health-Related Characteristics and mHealth App Use

Health status variables, including presence of heart conditions, lung disease, depression, smoking status, alcohol use, physical activity, and sleep patterns, were not significantly associated with mHealth app use after adjustment age, race/ethnicity, income, education level, and rural versus urban residence. None of the health-related variables demonstrated statistically significant associations with mHealth app use, suggesting that clinical status alone may not drive digital engagement among adults with hypertension and/or diabetes. (Table 4).

**Table 4:** Adjusted risk ratios for health app use for health-related factors adjusted for age, race/ethnicity, income, education, and rural/urban status

| Variable | Risk Ratio (95% CI) | p-value |
| --- | --- | --- |
| Condition |  |  |
| Has heart condition | 0.89 (0.66-1.22) | 0.458 |
| Has lung disease | 0.97 (0.77-1.22) | 0.795 |
| Has depression | 1.1 (0.89-1.34) | 0.354 |
| Smoking |  |  |
| Former | 1.04 (0.85-1.27) | 0.693 |
| Current | 0.8 (0.55-1.15) | 0.211 |
| Alcohol (drinks/day) | 1 (0.95-1.06) | 0.872 |
| Exercise (days/week) | 1.02 (0.99-1.06) | 0.227 |
| Sleep |  |  |
| Sleep (weekday hours) | 1.02 (0.94-1.1) | 0.632 |
| Sleep (weekend hours) | 0.99 (0.91-1.08) | 0.839 |

<sup>3</sup>**Note:** Reference categories were Smoking: *Never smoker*; Health condition variables: *No condition*; Continuous variables were modeled per unit increase.

## Discussion

This nationally representative analysis of U.S. adults with hypertension or diabetes reveals persistent disparities in mobile health app use. Despite widespread availability of digital health tools, engagement remains uneven across age, income, education, and digital access. These disparities reflect deeper structural gaps that shape chronic disease self-management and digital health adoption.

Age emerged as a key factor associated with mHealth app use, with older adults, particularly those aged 75 and above, reporting significantly lower engagement. Even after adjusting for socioeconomic status and technological access, the oldest age group had less than half the likelihood of app use compared to younger adults. This persistent gap suggests that barriers to digital health engagement extend beyond device ownership or internet access [24,25].

Generational differences in digital literacy, perceived utility, and trust in technology may contribute to lower uptake among older adults. These findings underscore the need for age-inclusive digital health strategies that prioritize usability, tailored training, and trust-building interventions to ensure equitable access across the lifespan. [26]. Prior work consistently documents lower mHealth uptake among older adults and highlights barriers that go beyond device ownership to include lower comfort with apps, sensory or cognitive challenges, and different perceptions of usefulness and trust [27,28]. These findings underscore the potential need for age-inclusive designs including simplified user interface and targeted digital onboarding strategies that accommodate cognitive, sensory, and motivational differences.

Socioeconomic status also played a critical role in mHealth app use [29]. Adults with annual incomes of $100,000 or more and those with postsecondary education were significantly more likely to use health apps. In contrast, lower-income groups showed no meaningful increases in usage, even when adjusting for smartphone ownership. This suggests that while affordability of devices may be necessary, it is not sufficient to drive engagement. In a nationwide study of racially diverse U.S. adults (N=1604) individuals with higher income and educational attainment were more likely to download and consistently use health-related apps. These findings point to broader economic privileges including stable internet access, smartphone ownership, digital literacy, and health-related self-efficacy as facilitators of digital empowerment. Such privilege enables users not only to access mHealth tools, but also to navigate their features confidently, trust their utility, and sustain meaningful engagement over time.

Digital access and confidence were significantly associated with mHealth app use, suggesting that disparities and self-efficacy may influence engagement with digital tools. Smartphone non-ownership and infrequent internet use were associated with dramatically lower odds of app engagement. Adults who never used the internet were 94 percent less likely to use health apps. Furthermore, individuals who reported frustration with technology or low confidence in using digital tools were significantly less engaged. A population-based study among German adults (n=1,500) with cardiovascular disease and diabetes showed that electronic health literacy was a key factor in health App use [30]. These findings reinforce the importance of digital literacy as a social determinant of health [31]. Tools for routine, rapid assessment of digital readiness and tailored support for patients who struggle with technology are needed in clinical settings.

Racial and ethnic disparities were not statistically significant in adjusted models. This contrasts with earlier studies that documented lower digital engagement among minoritized populations [32,33]. The absence of statistical significance may reflect evolving patterns of technology adoption or limitations in how race and ethnicity were measured in the HINTS dataset. It is important not to interpret this finding as evidence of equity achieved. Instead, it invites deeper exploration into intersectional barriers such as language, cultural relevance, and historical mistrust that may not be captured by broad racial categories. For example, while smartphone ownership is high across racial groups, Pew data show variation: 91 percent of White adults, 87 percent of Black adults, 93 percent of Hispanic adults, and 95 percent of Asian adults report owning smartphones. These differences, though narrowing, suggest that access alone may not fully explain disparities in digital health engagement [34].

Health-related variables, including comorbid conditions, smoking status, alcohol use, physical activity, and sleep, were not significantly associated with app use. This aligns with prior research showing that digital health engagement is often shaped by structural and psychosocial factors such as digital literacy, access to technology, usability, and social influence, rather than by clinical need [35,36]. The disconnect between disease burden and app use raises concerns about missed opportunities for intervention among those who might benefit most. It also highlights the need for proactive outreach and tailored digital solutions that resonate with patients lived experiences.

### Implications for practice and policy

Collectively, these findings illuminate the contours of the digital divide among people with hypertension and diabetes. They affirm the urgency of designing inclusive, culturally responsive mobile health interventions that address not only access but also usability, and relevance. We encourage clinicians to consider integrating digital literacy assessments into routine care of adults with hypertension and diabetes, and partner with community organizations to build digital capacity. Developers should prioritize co-design with underserved populations to ensure that tools are intuitive, meaningful, and accessible. Policymakers should invest in digital infrastructure and literacy programs that close the gap between availability and engagement.

Failure to address these disparities risks exacerbating existing inequities in chronic disease outcomes. As digital health becomes increasingly embedded in hypertension care, equity must be a guiding principle in design, implementation, and evaluation.

### Strengths and Limitations

This study offers several notable strengths. First, it utilizes data from the 2024 Health Information National Trends Survey (HINTS 7), a nationally representative dataset that enables generalizability of findings to U.S. adults with hypertension and/or diabetes. The inclusion of both English- and Spanish-speaking respondents enhances linguistic diversity and reflects real-world variation in digital health engagement. Second, this study applies a multidimensional framework that integrates sociodemographic, technological, and psychosocial factors, enabling a nuanced examination of the digital divide in chronic disease contexts. By considering multiple domains of access and engagement, the analysis moves beyond single-variable explanations and highlights structural disparities in mHealth adoption. Third, the use of survey-weighted log-binomial regression yields robust estimates of prevalence ratios while appropriately accounting for the complex sampling design, enhancing the generalizability and interpretability of findings.

However, several limitations should be acknowledged. The cross-sectional nature of the data precludes causal inference; observed associations may not reflect temporal relationships or directionality. All measures were self-reported, introducing potential for recall bias and social desirability bias, particularly in reporting app use and digital literacy. The outcome variable did not differentiate between types of mobile health applications (e.g., clinical vs. wellness-oriented), nor did it capture frequency or intensity of use. Additionally, unmeasured factors such as health literacy, language proficiency, and trust in digital platforms may influence engagement but were not assessed. Finally, the exclusion of non-English/Spanish speakers may underrepresent populations facing compounded barriers to digital health access.

## Conclusion

This study highlights persistent disparities in mobile health app use among U.S. adults with hypertension and diabetes, revealing that digital engagement is shaped more by structural and psychosocial factors than by clinical need. Age, income, education, and digital confidence emerged as key determinants, while health-related behaviors and comorbidities showed limited influence. These findings underscore the urgency of addressing the digital divide not only through expanded access but also through culturally responsive design, targeted literacy support, and inclusive implementation strategies.

As digital health tools become increasingly embedded in chronic disease management, equity must be central to their development and dissemination. Without intentional efforts to engage digitally marginalized populations, innovations in hypertension care risk reinforcing existing disparities. Future research should prioritize participatory approaches, intersectional analyses, and real-world evaluations to ensure that mobile health solutions are accessible, trusted, and effective for all.

## Conflict of Interest Disclosures**: None.**

### Author Contributions

All authors made significant contributions to the conception of the study, review of relevant articles, revising the manuscript, and critical revisions for important intellectual content. Each author provided final approval for the submitted version of the manuscript.

## Data Availability

The data that support the findings of this study are available from the corresponding author upon reasonable request.

## Acknowledgements

This research was, in part, funded by the National Institutes of Health (NIH) Agreement OT2HL158287. The views and conclusions contained in this document are those of the authors and should not be interpreted as representing the official policies, either expressed or implied, of the NIH. American Heart Association grants provided support for Nana Ofori Adomako (Grant Number: 25DIVSUP1474640).

## References

1. Hacker K. The Burden of Chronic Disease. Mayo Clin Proc Innov Qual Outcomes [Internet]. Elsevier; 2024 [cited 2025 Oct 11];8:112–9. 10.1016/J.MAYOCPIQO.2023.08.005

2. Martin SS, Aday AW, Allen NB, Almarzooq ZI, Anderson CAM, Arora P, et al. 2025 Heart Disease and Stroke Statistics: A Report of US and Global Data from the American Heart Association. Circulation. Lippincott Williams and Wilkins; 2025;151:e41–660. 10.1161/CIR.0000000000001303

1. Hill-Briggs F, Adler NE, Berkowitz SA, Chin MH, Gary-Webb TL, Navas-Acien A, et al. Social Determinants of Health and Diabetes: A Scientific Review. Diabetes Care [Internet]. American Diabetes Association Inc.; 2020 [cited 2025 Sep 12];44:258. 10.2337/DCI20-0053

2. Wang J, Tan F, Wang Z, Yu Y, Yang J, Wang Y, et al. Understanding Gaps in the Hypertension and Diabetes Care Cascade: Systematic Scoping Review. JMIR Public Health Surveill [Internet]. JMIR Publications Inc.; 2024 [cited 2025 Sep 12];10:e51802. 10.2196/51802

3. Mikulski BS, Bellei EA, Biduski D, De Marchi ACB. Mobile Health Applications and Medication Adherence of Patients With Hypertension: A Systematic Review and Meta-Analysis. Am J Prev Med [Internet]. Elsevier Inc.; 2022 [cited 2025 Oct 11];62:626–34. 10.1016/j.amepre.2021.11.003

4. de Souza Ferreira E, de Aguiar Franco F, dos Santos Lara MM, Levcovitz AA, Dias MA, Moreira TR, et al. The effectiveness of mobile application for monitoring diabetes mellitus and hypertension in the adult and elderly population: systematic review and meta-analysis. BMC Health Serv Res [Internet]. BioMed Central Ltd; 2023 [cited 2025 Oct 11];23:1–10. 10.1186/S12913-023-09879-6/FIGURES/4

5. Langford AT, Solid CA, Scott E, Lad M, Maayan E, Williams SK, et al. Mobile phone ownership, health apps, and tablet use in US adults with a self-reported history of hypertension: Cross-sectional study. JMIR Mhealth Uhealth [Internet]. JMIR Publications Inc.; 2019 [cited 2025 Oct 11];7:e12228. 10.2196/12228

6. Wang X, Wang B, Yin Tew W, Yang X, Xu X, Gao Y, et al. Exploring mHealth interventions for medication management: a scoping review of digital tools, implementation barriers, and patient outcomes. 10.7717/peerj-cs.3190

7. Fernández C, Vicente MA, Guilabert M, Carrillo I, Mira JJ. Developing a mobile health app for chronic illness management: Insights from focus groups. Digit Health [Internet]. SAGE Publications Inc.; 2023 [cited 2025 Sep 12];9:20552076231210664. 10.1177/20552076231210662

8. Ahmed MM, Okesanya OJ, Olaleke NO, Adigun OA, Adebayo UO, Oso TA, et al. Integrating Digital Health Innovations to Achieve Universal Health Coverage: Promoting Health Outcomes and Quality Through Global Public Health Equity. Healthcare [Internet]. Multidisciplinary Digital Publishing Institute (MDPI); 2025 [cited 2025 Sep 12];13:1060. 10.3390/HEALTHCARE13091060

9. Girmay M. Digital Health Divide: Opportunities for Reducing Health Disparities and Promoting Equitable Care for Maternal and Child Health Populations. International Journal of Maternal and Child Health and AIDS [Internet]. Scientific Scholar; 2024 [cited 2025 Sep 12];13:e026. 10.25259/IJMA_41_2024

10. Fitzpatrick PJ. Improving health literacy using the power of digital communications to achieve better health outcomes for patients and practitioners. Front Digit Health [Internet]. Frontiers Media SA; 2023 [cited 2025 Sep 12];5:1264780. 10.3389/FDGTH.2023.1264780

11. Lythreatis S, Singh SK, El-Kassar AN. The digital divide: A review and future research agenda. Technol Forecast Soc Change. Elsevier Inc.; 2022;175. 10.1016/J.TECHFORE.2021.121359

12. Yang R, Gao S, Jiang Y. Digital divide as a determinant of health in the U.S. older adults: prevalence, trends, and risk factors. BMC Geriatr [Internet]. BioMed Central Ltd; 2024 [cited 2025 Sep 12];24:1027. 10.1186/S12877-024-05612-Y

13. Tappen RM, Cooley ME, Luckmann R, Panday S. Digital Health Information Disparities in Older Adults: a Mixed Methods Study. J Racial Ethn Health Disparities [Internet]. Springer Science and Business Media Deutschland GmbH; 2021 [cited 2025 Sep 12];9:82. 10.1007/S40615-020-00931-3

14. del Pilar Arias López M, Ong BA, Frigola XB, Fernández AL, Hicklent RS, Obeles AJT, et al. Digital literacy as a new determinant of health: A scoping review. PLOS Digital Health [Internet]. Public Library of Science; 2023 [cited 2025 Sep 12];2:e0000279. 10.1371/JOURNAL.PDIG.0000279

15. van Kessel R, Wong BLH, Clemens T, Brand H. Digital health literacy as a super determinant of health: More than simply the sum of its parts. Internet Interv [Internet]. Elsevier B.V.; 2022 [cited 2025 Sep 12];27:100500. 10.1016/J.INVENT.2022.100500

16. Hoagland A, Kipping S. Challenges in Promoting Health Equity and Reducing Disparities in Access Across New and Established Technologies. Canadian Journal of Cardiology [Internet]. Elsevier; 2024 [cited 2025 Sep 12];40:1154–67. 10.1016/J.CJCA.2024.02.014

17. Saeed SA, Masters RMR. Disparities in Health Care and the Digital Divide. Curr Psychiatry Rep [Internet]. Springer; 2021 [cited 2025 Sep 12];23:61. 10.1007/S11920-021-01274-4

18. Health Information National Trends Survey | HINTS [Internet]. [cited 2025 Sep 12]. https://hints.cancer.gov/. Accessed 12 Sep 2025

19. Langford AT, Solid CA, Scott E, Lad M, Maayan E, Williams SK, et al. Mobile phone ownership, health apps, and tablet use in US adults with a self-reported history of hypertension: Cross-sectional study. JMIR Mhealth Uhealth [Internet]. JMIR Publications Inc.; 2019 [cited 2025 Oct 11];7:e12228. 10.2196/12228

20. Krebs P, Duncan DT. Health App Use Among US Mobile Phone Owners: A National Survey. JMIR Mhealth Uhealth [Internet]. JMIR mHealth and uHealth; 2015 [cited 2025 Oct 12];3:e101. 10.2196/mhealth.4924

21. Richardson DB, Kinlaw AC, MacLehose RF, Cole SR. Standardized binomial models for risk or prevalence ratios and differences. Int J Epidemiol [Internet]. Oxford University Press; 2015 [cited 2025 Sep 12];44:1660. 10.1093/IJE/DYV137

22. Yang R, Gao S, Jiang Y. Digital divide as a determinant of health in the U.S. older adults: prevalence, trends, and risk factors. BMC Geriatr [Internet]. BioMed Central Ltd; 2024 [cited 2025 Sep 30];24:1–11. 10.1186/S12877-024-05612-Y/TABLES/4

23. Wang K, Hernandez AM, Penate V, Abhat A, Casillas A. Digital Health Implementation Among Older Adults: Health Technology Navigators’ Perspectives. American Journal of Managed Care. Ascend Media; 2025;31:e125–31. 10.37765/AJMC.2025.89736

24. Finkelstein R, Wu Y, Brennan-Ing M. Older adults’ experiences with using information and communication technology and tech support services in New York City: findings and recommendations for post-pandemic digital pedagogy for older adults. Front Psychol [Internet]. Frontiers Media S.A.; 2023 [cited 2025 Sep 12];14:1129512. 10.3389/FPSYG.2023.1129512

25. Trinh M, Harris MT, Azevedo RFL, Rogers WA. Understanding older adults’ perceptions of mHealth apps. Gerontechnology. International Society for Gerontechnology; 2023;22. 10.4017/GT.2023.22.1.841.10

26. Bertolazzi A, Quaglia V, Bongelli R. Barriers and facilitators to health technology adoption by older adults with chronic diseases: an integrative systematic review. BMC Public Health [Internet]. BioMed Central Ltd; 2024 [cited 2025 Sep 12];24:506. 10.1186/S12889-024-18036-5

27. Liu P, Astudillo K, Velez D, Kelley L, Cobbs-Lomax D, Spatz ES. Use of Mobile Health Applications in Low-Income Populations. Circ Cardiovasc Qual Outcomes [Internet]. Lippincott Williams & WilkinsHagerstown, MD; 2020 [cited 2025 Sep 12];13:E007031. 10.1161/CIRCOUTCOMES.120.007031

28. Ernsting C, Stühmann LM, Dombrowski SU, Voigt-Antons J-N, Kuhlmey A, Gellert P. Associations of Health App Use and Perceived Effectiveness in People With Cardiovascular Diseases and Diabetes: Population-Based Survey. JMIR Mhealth Uhealth [Internet]. JMIR mHealth and uHealth; 2019 [cited 2025 Sep 12];7:e12179. 10.2196/12179

29. van Kessel R, Wong BLH, Clemens T, Brand H. Digital health literacy as a super determinant of health: More than simply the sum of its parts. Internet Interv [Internet]. Elsevier; 2022 [cited 2025 Sep 12];27:100500. 10.1016/J.INVENT.2022.100500

30. Ailawadhi S, Ailawadhi M, Dutta N, Parrondo RD, Roy V, Sher T, et al. The digital divide: Racial disparities in adoption and utilization of health information technology among patients with lymphoid cancers. Cancer Med [Internet]. John Wiley and Sons Inc; 2023 [cited 2025 Sep 12];12:19013. 10.1002/CAM4.6454

